# Development and psychometric properties of the Japanese Consumer Assessment of Healthcare Providers and Systems Clinician & Group Survey (CG-CAHPS)

**DOI:** 10.1101/2020.08.15.20175612

**Authors:** Takuya Aoki, Kuichiro Taguchi, Eiichi Hama

## Abstract

The Consumer Assessment of Healthcare Providers and Systems Clinician & Group Survey (CG-CAHPS) is one of the most widely studied and endorsed patient experience measures for ambulatory care. This study aimed to develop a Japanese CG-CAHPS and examine its psychometric properties. We evaluated the structural validity, criterion-related validity, and internal consistency reliability of the scale. Data were analyzed for 674 outpatients aged 18 years or older in 11 internal medicine clinics. The confirmatory factor analysis supported the scale’s structural validity and the same four composites (Access, Provider Communication, Care Coordination, and Office Staff) as that of the original CG-CAHPS. All provider-level Pearson correlation coefficients between the Japanese CG-CAHPS composites and overall provider rating exceeded the criteria. Results of item-total correlations and Cronbach’s alpha indicated adequate internal consistency reliability. We developed the Japanese CG-CAHPS as a valid and reliable scale to measure the quality of ambulatory care based on patient experience. The results of the Japanese CG-CAHPS survey will provide useful information to providers, organizations, and policy makers for achieving a patient-centered healthcare system in Japan.

## Introduction

The quality assessment of patient-centeredness from the patient’s perspective is an important aspect of quality of health care [1]. In recent years, better patient experience has been recognized as one of the crucial goals of healthcare alongside population health and per capita cost [2]. Patient experience is integrally tied to the principles and practices of patient- and family-centered care. Embedded within patient experience is a focus on individualized care and tailoring services to meet patients’ needs and engage them as partners in their care [3]. Numerous studies have shown that better patient experience is consistently associated with patient health outcomes, patient safety, and patient behaviors across a wide range of disease areas and settings [4–7].

The Consumer Assessment of Healthcare Providers and Systems Clinician & Group Survey (CG-CAHPS) is one of the most widely studied and endorsed patient experience measures for ambulatory care [8]. This standardized scale was developed by the Agency for Healthcare Research and Quality (AHRQ) and confirmed its validity and reliability [9,10]. Currently, in the United States, CG-CAHPS results are widely used as quality measures in accountability initiatives and to stimulate, guide, and monitor quality improvement efforts [11]. For example, results from the CG-CAHPS have been reported on the Physician Compare website, and insurers are also increasingly including patient experience data in pay-for-performance programs.

In contrast, in Japan, voluntary activities for the assessment of patient experience have just begun in limited settings, and systematic approaches for quality assessment and improvement based on patient experience measures are still unestablished. Only a few standardized scales, which have been confirmed psychometric properties, are available to assess patient experience in Japan [12–14]. Patient experience scales, which are tailored to different care settings, are needed to help identify aspects of care that can be targeted to improve patient experience. Accordingly, the present study aimed to develop a Japanese version of the CG-CAHPS and to examine its structural validity, criterion-related validity, and internal consistency reliability.

## Materials and methods

### Design, setting, and participants

The data used in this study were collected from a multicenter cross-sectional survey in 11 internal medicine clinics from February to March 2020. The 11 participating clinics voluntarily took part in the survey and are in urban areas in the Tokyo Metropolis and Kanagawa Prefecture, with all the clinics being privately owned and managed. In Japan, clinics are generally run by one full-time physician, nurses, and medical assistants, and they provide outpatient services and possibly home care. Independent surveyors distributed a self-administered questionnaire to all outpatients aged 18 years or older who visited one of the participating clinics within two days of the survey period using a continuous sampling method. Patients who were unable to respond to the questionnaire due to severe physical or mental disorders were excluded. Of the survey respondents, we excluded patients who visited the participating clinic for the first time during the survey period.

This study was approved by the Ethics Committee of Kyoto University Graduate School of Medicine (approval number R2331). Informed consent was not obtained because this study is an analysis of existing anonymous data.

## Measures

### CG-CAHPS

The original CG-CAHPS version 3.0 (the latest version as of February 2020) is an 18-item tool comprising four composites, a global rating, and five screening items (Q1, Q3, Q5, Q12, and Q15) [8]. The composites are Access (Q2, Q4, and Q6), Provider Communication (Q7, Q8, Q10, and Q11), Care Coordination (Q9, Q13, and Q16), and Office Staff (Q17 and Q18). The global rating is Rating of Provider (Q14).

Permission to translate the CG-CAHPS into Japanese was granted by the AHRQ. Following the translation guidelines for CAHPS® surveys provided by AHRQ [15], translation of the CG-CAHPS into Japanese was performed through the following steps. First, two bilingual translators with experience in translating survey instruments conducted forward translations from English to Japanese. Two forward translations were performed independently. The two translations were then reviewed by a translation reviewer who is a native speaker of Japanese and has experience in translating survey instruments. After reviewing the translation, the reviewer prepared a reconciled version of the translation. A committee consisting of the two translators and the reviewer then discussed and prepared the final version. Revisions were made to the reconciled version necessary for cross-cultural adaptation. The final wording of each survey item and response option was determined by consensus.

The CG-CAHPS survey uses multiple response formats: four-point Likert scales (1 = never, 2 = sometimes, 3 = usually, and 4 = always), and a global rating scale (0 = worst to 10 = best). To make the results easier to understand, we converted all scales to normalized scores ranging from 0 to 100 using the following formula:

Normalized Score = 100 * (Respondent’s selected response value – Minimum response value on the scale) / (Maximum response value – Minimum response value)

In the Japanese version, assuming the convergence in each composite as in the original version, the score for each of the four composites was computed as the mean value for all normalized scores in the scale that would fall in the range of 0–100 points, with higher scores indicating better performance.

### Statistical analysis

We validated the Japanese CG-CAHPS through the following steps:

First, we carried out a confirmatory factor analysis to evaluate the structural validity of the Japanese CG-CAHPS composites. In the factor analysis, we hypothesized the same factor structure (four-factor solution) as that of the original CG-CAHPS. The appropriateness of the resulting structure was determined by examining if factor loadings were 0.40 or greater [16]. Model fitness was assessed using the goodness-of-fit index (GFI), comparative fit index (CFI), root mean square error of approximation (RMSEA), and standardized root mean square residual (SRMR). For GFI and CFI, a value of > 0.90 is considered acceptable, and a value of > 0.95 indicates excellent goodness of fit. Previous studies suggest that models with RMSEA < 0.07 and SRMR < 0.08 are representative of models with a good fit [17–19].

Second, we used the Japanese CG-CAHPS composite scores and the overall provider rating to examine criterion-related validity. Validity was assessed using Pearson correlation coefficients with each Japanese CG-CAHPS composite to predict the Rating of Provider (0 = Worst to 10 = Best) of the scale at the provider-level. A correlation coefficient greater than 0.30 was considered meaningful [20]. Provider-level correlations are a more important criterion for measurement than patient-level correlations because the former are benchmarking tools to compare one provider or facility with another. To examine provider-level correlations, we used each provider’s mean score on CG-CAHPS composites and the Rating of Provider.

Internal consistency reliability was examined by item-total correlations and Cronbach’s alpha. For a scale to be considered sufficiently reliable, an item-total correlation of 0.30 and a Cronbach’s alpha value of 0.70 is recommended [21]. Finally, descriptive statistics were performed on the Japanese CG-CAHPS scores, including the mean, standard deviation, and observed range. To deal with missing data, in the confirmatory factor analysis, we used the full information maximum likelihood estimation to enable the use of information collected from participants with missing data. In the evaluation of criterion-related validity and internal consistency, we conducted complete case analyses. All statistical analyses were conducted using R version 3.6.3 (R Foundation for Statistical Computing, Vienna, Austria; https://www.R-project.org).

## Results

Of the total 818 eligible outpatients, 787 (96.2%) responded to the survey. Of these respondents, we excluded 113 who visited the participating clinic for the first time and based analyses on the remaining 674 patients. Table 1 shows the participants’ characteristics.

**Table 1:** Participants’ characteristics (N = 674)

| Characteristic | n (%) |
| --- | --- |
| Gender |  |
| Male | 251 (37.9) |
| Female | 411 (62.0) |
| Data missing | 12 |
| Age (years) |  |
| 18–24 | 34 (5.1) |
| 25–34 | 125 (18.8) |
| 35–44 | 114 (17.2) |
| 45–54 | 126 (19.0) |
| 55–64 | 109 (16.4) |
| 65–74 | 86 (13.0) |
| 75 or more | 70 (10.5) |
| Data missing | 10 |
| Education |  |
| Less than high school | 34 (5.1) |
| High school | 192 (29.0) |
| Junior college | 136 (20.5) |
| More than or equal to college | 301 (45.4) |
| Data missing | 11 |
| Self-rated Health |  |
| Excellent | 19 (2.9) |
| Very good | 77 (11.6) |
| Good | 160 (24.1) |
| Fair | 317 (47.7) |
| Poor | 92 (13.8) |
| Data missing | 9 |
| Duration of relationship with provider |  |
| Less than 6 months | 222 (33.3) |
| At least 6 months but less than 1 year | 116 (17.4) |
| At least 1 year but less than 3 years | 147 (22.0) |
| At least 3 years but less than 5 years | 66 (9.9) |
| 5 years or more | 116 (17.4) |
| Data missing | 7 |

Table 2 shows the participants’ responses to each item of the Japanese CG-CAHPS. The Top Box score for each item, which is the percentage of participants who provided the most positive responses on that item, ranged from 56.1% to 73.6%. Regarding the mean Top Box score for composites, the highest score was observed for Provider Communication (70.9%), while the lowest score was for Care Coordination (59.6%). The bottom box score, which is the percentage of participants with the least positive responses on the item, ranged from 0.8% to 7.2%.

**Table 2:** Response to Japanese CG-CAHPS items (N = 674): number (%)

|  | Never | Sometimes | Usually | Always | Data missing | Not applicable<sup>a</sup> |
| --- | --- | --- | --- | --- | --- | --- |
| Access: |  |  |  |  |  |  |
| Q2. In the last 6 months, when you contacted this provider's office to get an appointment for care you needed right away, how often did you get an appointment as soon as you needed? | 7 (3.6) | 16 (8.3) | 45 (23.4) | 124 (64.6) | 10 | 472 |
| Q4. In the last 6 months, when you made an appointment for a check-up or routine care with this provider, how often did you get an appointment as soon as you needed? | 11 (4.5) | 27 (11.1) | 43 (17.6) | 163 (66.8) | 12 | 418 |
| Q6. In the last 6 months, when you contacted this provider's office during regular office hours, how often did you get an answer to your medical question that same day? | 5 (3.4) | 16 (10.7) | 32 (21.5) | 96 (64.4) | 0 | 525 |
| Provider Communication: |  |  |  |  |  |  |
| Q7. In the last 6 months, how often did this provider explain things in a way that was easy to understand? | 6 (0.9) | 27 (4.1) | 139 (21.3) | 480 (73.6) | 22 | – |
| Q8. In the last 6 months, how often did this provider listen carefully to you? | 8 (1.2) | 25 (3.8) | 148 (22.4) | 480 (72.6) | 13 | – |
| Q10. In the last 6 months, how often did this provider show respect for what you had to say? | 8 (1.2) | 25 (3.8) | 148 (22.7) | 471 (72.2) | 22 | – |
| Q11. In the last 6 months, how often did this provider spend enough time with you? | 8 (1.2) | 41 (6.3) | 178 (27.3) | 426 (65.2) | 21 | – |
| Care Coordination: |  |  |  |  |  |  |
| Q9. In the last 6 months, how often did this provider seem to know the important information about your medical history? | 31 (4.7) | 40 (6.1) | 205 (31.4) | 377 (57.7) | 21 | – |
| Q13. In the last 6 months, when this provider ordered a blood test, x-ray, or other test for you, how often did someone from this provider's office follow up to give you those results? | 24 (7.2) | 24 (7.2) | 69 (20.7) | 216 (64.9) | 15 | 326 |
| Q16. In the last 6 months, how often did you and someone from this provider's office talk about all the prescription medicines you were taking? | 34 (5.9) | 73 (12.6) | 147 (25.4) | 324 (56.1) | 20 | 76 |
| Office Staff: |  |  |  |  |  |  |
| Q17. In the last 6 months, how often were clerks and receptionists at this provider's office as helpful as you thought they should be? | 9 (1.4) | 49 (7.5) | 214 (32.7) | 383 (58.5) | 19 | – |
| Q18. In the last 6 months, how often did clerks and receptionists at this provider's office treat you with courtesy and respect? | 5 (0.8) | 43 (6.6) | 182 (27.8) | 425 (64.9) | 19 | – |
|  | 0–2 | 3–5 | 6–8 | 9–10 | Data missing | Not applicable<sup>a</sup> |
| Rating of Provider: |  |  |  |  |  |  |
| Q14. Using any number from 0 to 10, where 0 is the worst provider possible and 10 is the best provider possible, what number would you use to rate this provider? | 3 (0.5) | 33 (5.1) | 204 (31.2) | 414 (63.3) | 20 | – |

<sup>a</sup>The number of participants who skipped the item due to the response to the screenig item.

### Structural validity

Fig 1 shows the path diagrams of the confirmatory factor analysis to assess the structural validity of the Japanese CG-CAHPS composites. All factor loadings of each item onto each factor were above the 0.40 criteria, ranging from 0.48 to 0.90. The correlation coefficients among factors ranged from 0.51 to 0.89. The conceptual model indicated good fit, with GFI = 0.988, CFI =0.940, and SRMR = 0.067. However, the RMSEA was 0.074, slightly above the cutoff at 0.07.

**Figure 1:**
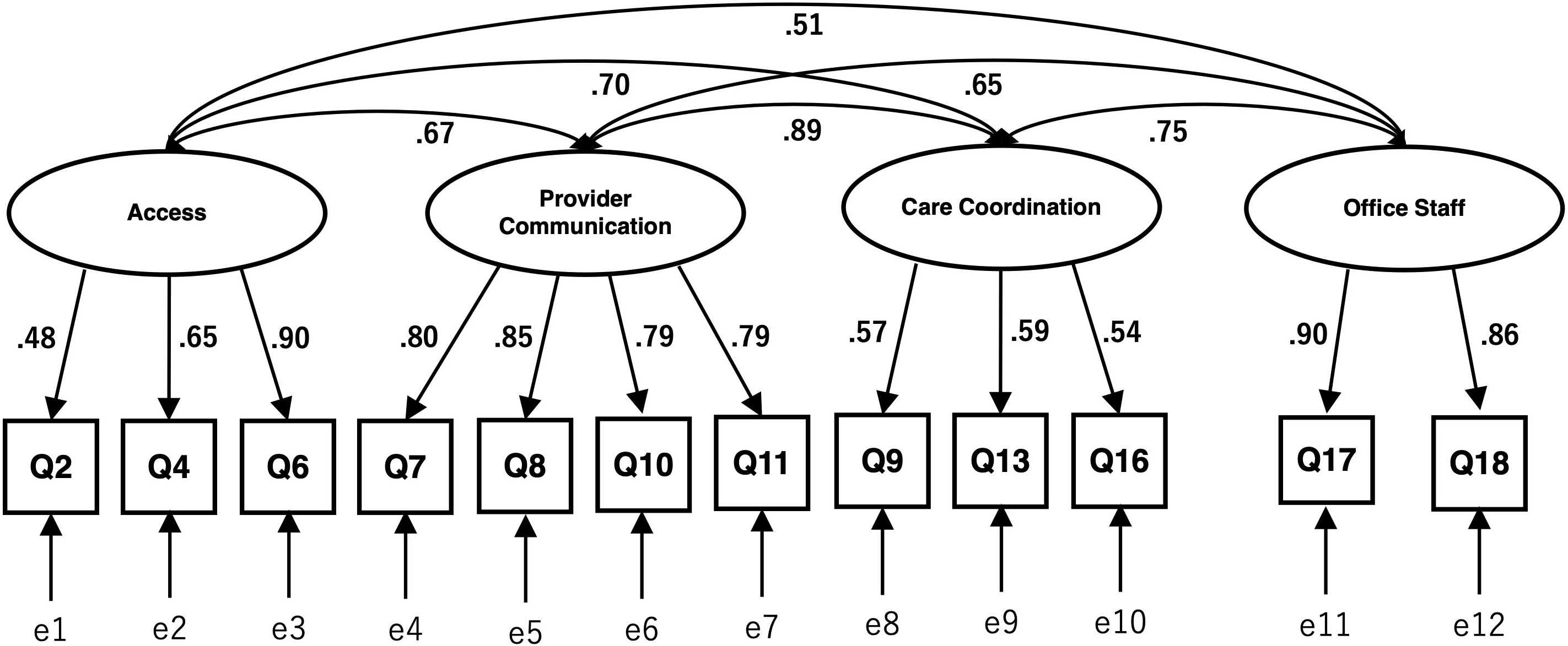
Factor structure of Japanese CG-CAHPS (confirmatory factor analysis) Squares are observed variables (items); ellipses are latent variables (factors), values on the single-headed arrows are standardized factor loadings, values on the double-headed arrows are correlation coefficients. CG-CAHPS = Consumer Assessment of Healthcare Providers and Systems Clinician & Group Survey.

### Criterion-related validity

Table 3 shows the Pearson correlation coefficients between the Japanese CG-CAHPS composites and the Rating of Provider as an overall provider rating at the provider-level. All correlations exceeded the 0.30 criterion. Provider Communication (*r* = 0.85) had the highest correlation with the overall rating.

**Table 3:** Pearson correlation coefficients between Japanese CG-CAHPS composites and overall provider rating

| Composites | Provider-level correlations |
| --- | --- |
| Access | 0.59 |
| Provider Communication | 0.85 |
| Care Coordination | 0.79 |
| Office Staff | 0.49 |

### Internal consistency reliability and descriptive statistics

Table 4 indicates the score distribution and internal consistency reliability for the Japanese CG-CAHPS. All item-total correlations were above the 0.30 criteria, ranging from 0.31 to 0.92. For Access, Provider Communication, and Office Staff, the Cronbach’s alpha was above 0.70. However, for Care Coordination, the Cronbach’s alpha did not exceed the 0.70 criterion.

Descriptive statistics showed that the highest scored scale was Provider Communication (mean score = 88.1), and the most poorly scored scale was Care Coordination (mean score = 78.6). The full range of possible scores was observed for all scales.

**Table 4:** Descriptive features and internal consistency reliability of Japanese CG-CAHPS (N = 674)

|  | Number of items | Mean | Standard deviation | Observed range | Item-total correlation | Cronbach's alpha |
| --- | --- | --- | --- | --- | --- | --- |
| Composites |  |  |  |  |  |  |
| Access | 3 | 83.9 | 25.0 | 0.0–100.0 | 0.53–0.92 | 0.81 |
| Provider Communication | 4 | 88.1 | 17.8 | 0.0–100.0 | 0.73–0.78 | 0.88 |
| Care Coordination | 3 | 78.6 | 23.3 | 0.0–100.0 | 0.31–0.44 | 0.58 |
| Office Staff | 2 | 84.2 | 21.0 | 0.0–100.0 | 0.77 | 0.87 |
| Global ratings |  |  |  |  |  |  |
| Rating of Provider | 1 | 87.9 | 15.6 | 0.0–100.0 | – | – |

## Discussion

We developed the Japanese CG-CAHPS in the form of a standardized scale for assessing the quality of ambulatory care from the patient’s perspective in Japan. In our multicenter study, the psychometric properties of the Japanese CG-CAHPS, including structural validity, criterion-related validity, and internal consistency reliability, were evaluated. Even in Japan, it is important to have valid and reliable measures for assessing patient experience in various settings.This scale could be used for quality improvement based on the assessment of patient experience with ambulatory care and for health services research in Japan.

The confirmatory factor analysis supported the scale’s structural validity and the same four composites (Access, Provider Communication, Care Coordination, and Office Staff) as that of the original CG-CAHPS. Correlation coefficients between all Japanese CG-CAHPS composites and the overall provider rating for assessing criterion-related validity exceeded the meaningful value at the provider-level. In internal consistency analyses, only Cronbach’s alpha for Care Coordination did not exceed the recommended value. Cronbach’s alpha is quite sensitive to the number of items in the scale; therefore, it is common to find low Cronbach’s alpha for scales with few items [22]. In this case, it is more appropriate to report the item-total or inter-item correlation. In our study, all item-total correlations were greater than the cutoff value, which indicated acceptable internal consistency of the scales.

The CG-CAHPS is one of the most widely studied patient experience scales for ambulatory care worldwide. The CG-CAHPS has been translated into many languages in order to be used in other countries so that comparisons of health service quality from the patient perspective can be made. In our study, the recovery rate for the questionnaire administered was very high, suggesting a low risk of selection bias.

However, the present study has several potential limitations. First, in this study, we evaluated the structural validity, criterion-related validity, and internal consistency reliability of the Japanese CG-CAHPS, other psychometric properties, including convergent and discriminant validity, test-retest reliability, and interpretability, have not been assessed [23]. These psychometric properties of the scale need to be evaluated in future studies. Second, our survey setting was restricted to urban areas and may not have sufficiently represented the Japanese national level. Therefore, the study results may have limited generalizability and a survey using the Japanese CG-CAHPS in other suburban and rural areas should be conducted.

## Conclusions

We developed the Japanese CG-CAHPS as a valid and reliable scale to measure the quality of ambulatory care based on patient experience. The results of the Japanese CG-CAHPS survey will provide useful information to providers, organizations, and policy makers for achieving a patient-centered healthcare system in Japan.

## Data Availability

Data cannot be shared publicly because additional unpublished data is still being analyzed for another research and only available to the members of the study team.

